# COVID-19 outcomes, risk factors and associations by race: a comprehensive analysis using electronic health records data in Michigan Medicine

**DOI:** 10.1101/2020.06.16.20133140

**Authors:** Tian Gu, Jasmine A. Mack, Maxwell Salvatore, Swaraaj Prabhu Sankar, Thomas S. Valley, Karandeep Singh, Brahmajee K. Nallamothu, Sachin Kheterpal, Lynda Lisabeth, Lars G. Fritsche, Bhramar Mukherjee

## Abstract

**Importance:** Blacks/African-Americans are overrepresented in the number of COVID-19 infections, hospitalizations and deaths. Reasons for this disparity have not been well-characterized but may be due to underlying comorbidities or sociodemographic factors.

**Objective:** To systematically determine patient characteristics associated with racial/ethnic disparities in COVID-19 outcomes.

**Design:** A retrospective cohort study with comparative control groups.

**Setting:** Patients tested for COVID-19 at University of Michigan Medicine from March 10, 2020 to April 22, 2020.

**Participants:** 5,698 tested patients and two sets of comparison groups who were not tested for COVID-19: randomly selected unmatched controls (n = 7,211) and frequency-matched controls by race, age, and sex (n = 13,351).

**Main Outcomes and Measures:** We identified factors associated with testing and testing positive for COVID-19, being hospitalized, requiring intensive care unit (ICU) admission, and mortality (in/out-patient during the time frame). Factors included race/ethnicity, age, smoking, alcohol consumption, healthcare utilization, and residential-level socioeconomic characteristics (SES; i.e., education, unemployment, population density, and poverty rate). Medical comorbidities were defined from the International Classification of Diseases (ICD) codes, and were aggregated into a comorbidity score.

**Results:** Of 5,698 patients, (median age, 47 years; 38% male; mean BMI, 30.1), the majority were non-Hispanic Whites (NHW, 59.2%) and non-Hispanic Black/African-Americans (NHAA, 17.2%). Among 1,119 diagnosed, there were 41.2% NHW and 37.4% NHAA; 44.8% hospitalized, 20.6% admitted to ICU, and 3.8% died. Adjusting for age, sex, and SES, NHAA were 1.66 times more likely to be hospitalized (95% CI, 1.09-2.52; *P=*.02), 1.52 times more likely to enter ICU (95% CI, 0.92-2.52; *P*=.10). In addition to older age, male sex and obesity, high population density neighborhood (OR, 1.27 associated with one SD change [95% CI, 1.20-1.76]; *P*=.02) was associated with hospitalization. Pre-existing kidney disease led to 2.55 times higher risk of hospitalization (95% CI, 1.62-4.02; *P*<.001) in the overall population and 11.9 times higher mortality risk in NHAA (95% CI, 2.2-64.7, *P*=.004).

**Conclusions and Relevance:** Pre-existing type II diabetes/kidney diseases and living in high population density areas were associated with high risk for COVID-19 susceptibility and poor prognosis. Association of risk factors with COVID-19 outcomes differed by race. NHAA patients were disproportionately affected by obesity and kidney disease.

**Key Points:** *Question:* What are the sociodemographic and pre-existing health conditions associated with COVID-19 outcomes and how do they differ by race/ethnicity?

*Findings:* In this retrospective cohort of 5,698 patients tested for COVID-19, high population density and comorbidities such as type II diabetes/kidney disease were associated with hospitalization, in addition to older age, male sex and obesity. Adjusting for covariates, non-Hispanic Blacks were 1.66 times more likely to be hospitalized and 1.52 times more likely to be admitted to ICUs than non-Hispanic Whites.

*Meaning:* Targeted interventions to support vulnerable populations are needed. Racial disparities existed in COVID-19 outcomes that cannot be explained after controlling for age, sex, and socioeconomic status.

## Introduction

The COVID-19 pandemic, caused by severe acute respiratory syndrome coronavirus SARS-CoV-2, has demonstrated racial disparities in those affected in the United States (US)^1–13^. In the state of Michigan in particular, there have been 64,998 confirmed COVID-19 cases and 5,943 deaths as of June 11, 2020, which makes Michigan one of the most affected states in the US^14^. While Blacks/African-Americans represent 14% of the Michigan population^15^, they account for 31% of COVID-19 cases and 40% of deaths attributed to COVID-19^14^. Similar trends are observed in New York^9^ and Illinois, where there is an overrepresentation of African-Americans and Latinos in COVID-19 cases and deaths^16^.

Overrepresentation of minority populations in poorer COVID-19 outcomes may be explained by a myriad of factors, such as by weathering, or early health deterioration due to cumulative impact of socioeconomic disparity ^17,18^, higher comorbidity burden^19^, inadequate healthcare^19^, and socioeconomic differences related to unemployment, food insecurity, and housing instability^17^. Several studies have reported non-White, male, older age, current smoking, and comorbid conditions as high risk factors of COVID-19 susceptibility and hospitalization^2,13,20–24^. Racial/ethnic minorities who maintain livelihood as essential workers are more likely to be exposed to the virus^16^, whereas living in high density areas^1^, high proportion of homelessness^25^ and incarceration^26^ adds to the barriers to social distancing^16^.

Although studies had reported many possible reasons for the overrepresentation of minority populations in poorer health outcomes, the evidence supporting the observed disparity in COVID-19 outcomes remains limited, and more data from diverse communities need to be analyzed. In addition, experiences from COVID-19 highlight the need to not only identify risk factors but also to avoid spurious conclusions of racial/ethnic differences being explained by biology, which could further perpetuate racial/ethnic stereotypes^17^. Data on holistic clinical and sociodemographic factors contributing to racial/ethnic differences in COVID-19 outcomes is limited. Some previous studies have also compared those who tested positive for COVID-19 to those who are negative, instead of population-based controls where selection bias is potentially observed^27,28^.

The objective of this study is to determine sociodemographic and comorbid conditions that are associated with COVID-19 outcomes (e.g., testing positive, hospitalization, admission to ICU, and mortality), utilizing electronic health records (EHR) from the University of Michigan, which serves a large patient population in the US Midwest.

## Subjects and Methods

### Evaluation cohorts

#### COVID-19 cohort

We extracted the EHR data for patients tested for COVID-19 at the University of Michigan Medicine Health System, also known as Michigan Medicine (MM), from March 10, 2020 to April 22, 2020. Our study cohort of 5,698 patients comprises 5,500 patients (96.5%) who were tested at MM and 198 patients (3.5%) who were treated for COVID-19 in MM but tested elsewhere, of which 1,119 were COVID-19 positive. For ease of notation, we refer to them as **the tested cohort (n=5**,**698)** and **the positive cohort (n=1**,**119)**. The tested cohort is a non-random sample of the population, since the testing protocol at MM focused on prioritized testing^29^ (e.g., testing symptomatic patients and those at the highest risk of exposure). This cohort also contained transfer patients from other hospitals.

#### Control selection

To understand how selection bias factored into our sample, in addition to comparing COVID-19 positive patients with those testing negative, we created two sets of controls from the MM database. The first **unmatched control group (n=7**,**211)** is a similar-sized random sample of contemporaneous patients. The second 1:3 **frequency-matched control group (n=13**,**351)** is matched by race, sex and age (above or below 50). All controls were alive at the time of data extraction. Study protocols were reviewed and approved by the University of Michigan Medical School Institutional Review Board (IRB ID HUM00180294 and HUM00155849).

#### Description of variables

A summary data dictionary, eTable S2A, is available in Supplement with source and definition of each variable used in our analysis.

#### COVID-19 prognosis outcomes

Among the patients diagnosed with COVID-19, we considered various stages of progression of the disease that included hospitalization, admission to the ICU and death. Hospitalizations were defined by inpatients with a COVID-19 diagnosis where the admission date was within the time frame of the data extraction. ICU patients were defined as patients who were admitted to ICU units any time during their COVID-19 related hospitalization. Mortality data including inpatient and non-hospitalized deaths was extracted from EHR.

#### Classifying patients who were still in hospital and ICU

We categorized patients into non-hospitalized, hospitalized (includes ICU stays), and hospitalized with ICU stay based on the admission and discharge data. Several patients were still admitted in the hospital (non-ICU, n=53) or were still in an ICU (n=113) at the time of the data extraction. We performed a sensitivity analysis by excluding these patients whose final prognostic outcome is unclear from the analysis (eTable S4 in Supplement).

#### Generation of comorbidities from electronic health records

We constructed the comorbid conditions using available International Classification of Diseases (ICD; ninth and tenth editions) code for 23,769 individuals (n_tested_: 5,225, n_unmatched_: 6,811, n_matched_: 11,733) from EHR. Longitudinal time-stamped diagnoses were recoded to indicator variables for whether a patient ever had a given diagnosis code recorded by MM. To differentiate *pre-existing* conditions from diagnoses related to COVID-19 testing/treatment, we applied a 14-day-prior restriction on the tested cohort by removing diagnoses that first appeared within the 14 days before the first test or diagnosis date, whichever was earlier (4,622 of the 5,225 tested individuals had diagnoses data after the 14-day-prior restriction). We focused on seven binary disease indicators that have been specifically mentioned in relation to COVID-19 outcomes: respiratory, circulatory, any cancer, type II diabetes, kidney, liver, and autoimmune diseases (ICD codes in eTable S2A in Supplement). We calculated a comorbidity score as the sum of these seven that ranges from 0-7. For exploratory analysis, we defined prior medication use as at least one appearance of a given class of medication in the patient’s EHR.

#### Defining race/ethnicity groups, SES and other adjustment covariates

Variables such as self-reported sex, race/ethnicity, smoking status, alcohol consumption, body mass index (BMI), and age were extracted from the EHR. We classified a patient to be seeking primary care in MM if they have had an encounter in any of the primary care locations in MM since January 1, 2018. Measures of socioeconomic characteristics are defined by US census tract (based on residential address available in each patient’s EHR) for the year 2010. The boundaries for the census tracts were normalized to 2010 tract boundaries using the Longitudinal Tract Data Base ^30^. We chose three SES indicators included in the National Neighborhood Data Archive (NaNDA)^31^: percentage of population with below high school (<HS) education, percentage unemployed and percentage with annual income below federal poverty level (FPL). We also used population density per square mile as a potential predictor^31^.

### Statistical analysis

Since all outcomes were binary, we performed logistic regression to assess the risk factors of COVID-19 outcomes, by reporting Firth bias-corrected estimate of the odds ratio to address potential separation issues, as well as 95% Wald-type confidence interval and *P-value*. Continuous predictors were standardized before modeling. Three sets of adjustment covariates were used to check the robustness of inference to the choice of potential confounders: (i) age, race/ethnicity, sex, (ii) adjustment (i)+SES, and (iii) adjustment (ii)+comorbidity score. We present the results of adjustment (ii) in the main text, since adjustment (iii) led to loss of sample size as many of the patients did not have EHR data to construct the pre-existing conditions. Sensitivity analysis after removing those still in ICU or hospital at the time of data extraction were also performed (eTable S4 in Supplement).

Three different types of association models were created (eTable S2B in Supplement):

a. <u>Predictors of being tested:</u> comparing the tested cohort with those who were not tested for COVID-19 (unmatched controls).
b. <u>Predictors of COVID-19 susceptibility:</u> comparing the positive cohort with those who were not diagnosed with COVID-19 (unmatched controls)
c. <u>Predictors of three COVID-19 prognostic outcomes:</u> among the positive cohort, comparing those who were hospitalized with those who were not, (ii) those who were admitted to ICU with those who were not, and (iii) those who died with those who did not. (this analysis does not use untested controls)

After analyzing the full cohort including all races, we carried out association analysis stratified by race/ethnicity (NHW or NHAA, given limited sample size for other racial/ethnic groups). Frequency-matched, instead of unmatched controls, were used to assess the COVID-19 susceptibility as the proportion of race/ethnicity in unmatched controls are not comparable to the stratified study population.

Analyses were performed in R version 3.6.2 (R Foundation for Statistical Computing). Statistical significance was defined using a 2-sided significance level of α□=□.05. All unadjusted models and results using other adjustments can be found in eTable S3A for all cohort, S3B and S3C for race/ethnicity-stratified in Supplement.

## Results

### Descriptive Statistics

A total of 5,698 patients were tested for COVID-19 (median age, 47 years [IQR, 31]; 38% male [2030/5336]; mean BMI 30.1 [SD, 8.1]), among whom 1,119 (19.6%) tested positive (**Table 1**). Over half of the tested (3,026, 53.1%) had primary care at MM. A majority of those tested were either NHW (3,374, 59.2%) or NHAA (981, 17.2%). Among the 1,119 positive patients, 44.8% (501) were hospitalized, 20.6% (231) were in an ICU, and 3.8% (43) died. As the disease progressed among patients testing positive (from non-hospitalized to hospitalized, ICU, and deceased), the proportion of older age (65 and above), male sex, and ever-smoker, consistently increased (**Table 1**). BMI showed an increasing trend from non-hospitalized to hospitalized and ICU but not in those who died (pairwise comparison results in eTable S1B in Supplement). The descriptive trends of all three residential SES variables indicate lower SES is associated with poorer COVID outcomes (**Table 1**). Both the tested cohort and the positive cohort had a higher enrichment of diseases. In particular, type II diabetes and kidney diseases were more common in groups with more severe outcomes. The tested population were also more frequent users of medication (e.g. NSAIDs, statins, and anticoagulants) compared with the unmatched controls (eTable S1A in Supplement). Descriptive statistics stratified by NHW and NHAA (eTable S1A and S1B in Supplement) show a stark contrast in COVID-19 outcomes across these two groups that we capture in **Figure 1**. The test-positive rate was significantly higher in NHAA compared with NHW (42.6% vs. 13.7%, *P*<.001), hospital admission (52.2% vs. 39%, *P*<.001), ICU (27% vs. 14.8%, *P*<.001), and death (5.3% vs. 3.0%, *P*=.12). Moreover, racial disparities also exist across demographics such as BMI, lower SES, and comorbidity conditions, which were consistently more prevalent in NHAA compared with NHW across subpopulations defined by COVID-19 outcomes (pairwise *P*<.001, eTable S1C in Supplement).

**Table 1.** Descriptive Characteristics of the COVID-19 Tested/Diagnosed cohort

| Variable | COVID-19 Tested |  | COVID-19 Positive |  |  |  |
| --- | --- | --- | --- | --- | --- | --- |
|  | Overall<br>(n=5698) | Negative<br>(n=4579) | Overall<br>(n=1119) | Hospitalized<br>(n=501) | ICU<br>(n=231) | Deceased<br>(n=43) |
|  | No./No. (%) |  |  |  |  |  |
| **Age (in years)** |  |  |  |  |  |  |
| mean (SD); n | 47.3 (20.9); 5698 | 45.9 (21.3); 4579 | 52.8 (17.9); 1119 | 60.9 (16.3); 501 | 59.2 (15.8); 231 | 68.3 (12.8); 43 |
| median (IQR) | 47 (31) | 45 (31) | 53 (28) | 63 (22) | 62 (19) | 70 (17) |
| **Age Range, y** |  |  |  |  |  |  |
| [0,18) | 374 (6.56) | 365 (7.97) | 9 (0.8) | 5 (0.81) | 4 (0.8) | 3 (1.3) |
| [18,35) | 1313 (23.04) | 1109 (24.22) | 204 (18.23) | 171 (27.67) | 33 (6.59) | 19 (8.23) |
| [35,50) | 1358 (23.83) | 1101 (24.04) | 257 (22.97) | 177 (28.64) | 80 (15.97) | 33 (14.29) |
| [50,65) | 1377 (24.17) | 1040 (22.71) | 337 (30.12) | 172 (27.83) | 165 (32.93) | 82 (35.5) |
| [65,80) | 923 (16.2) | 699 (15.27) | 224 (20.02) | 77 (12.46) | 147 (29.34) | 75 (32.47) |
| [80,100) | 353 (6.2) | 265 (5.79) | 88 (7.86) | 16 (2.59) | 72 (14.37) | 19 (8.23) |
| **Male Sex** | 2030/5336 (38) | 1524/4246 (35.9) | 506/1090 (46.4) | 280/496 (56.5) | 138/227 (60.8) | 28/43 (65.1) |
| **Primary Care in MM** | 3026/5698 (53.1) | 2486/4579 (54.3) | 540/1119 (48.3) | 127/501 (25.3) | 45/231 (19.5) | 7/43 (16.3) |
| **BMI, mean (SD); No.** | 30.1 (8.1); 4608 | 29.4 (7.5); 3609 | 32.4 (9.5); 999 | 33.6 (11.1); 467 | 35.3 (13.3); 217 | 31.5 (8.0); 42 |
| **BMI Range, y** |  |  |  |  |  |  |
| <18.5 | 76/4608 (1.3) | 68/3609 (1.5) | 8/999 (0.7) | 4/467 (0.8) | 1/217 (0.4) | 1/42 (2.3) |
| [18.5, 25) | 196/4608 (21) | 1033/3609 (22.6) | 163/999 (14.6) | 55/467 (11) | 23/217 (10) | 6/42 (14) |
| [25, 30) | 1394/4608 (24.5) | 1091/3609 (23.8) | 303/999 (46.9) | 150/467 (29.9) | 60/217 (26) | 15/42 (34.9) |
| >=30 | 1942/4608 (34.1) | 1417/3609 (30.9) | 525/999 (10.7) | 258/467 (51.5) | 133/217 (57.6) | 20/42 (46.5) |
| **Ever-Smoker** | 1806/4699 (38.4) | 1502/3815 (39.4) | 304/884 (34.4) | 148/373 (39.7) | 64/134 (47.8) | 14/20 (70) |
| **Smoking Status, No. (%)** |  |  |  |  |  |  |
| Never-Smoker | 2893/4699 (61.6) | 2313/3815 (60.6) | 580/884 (65.6) | 225/373 (60.3) | 70/134 (52.2) | 6/20 (30) |
| Past-Smoker | 1406/4699 (29.9) | 1142/3815 (29.9) | 264/884 (29.9) | 138/373 (37) | 61/134 (45.5) | 14/20 (70) |
| Current-Smoker | 400/4699 (8.5) | 360/3815 (9.4) | 40/884 (4.52) | 10/373 (2.7) | 3/134 (2.2) | 0/20 (0) |
| **Alcohol Consumption** | 2573/3879 (66.3) | 2134/3198 (66.7) | 439/681 (64.5) | 151/243 (62.1) | 61/91 (67) | 13/16 (81.2) |
| **Race Ethnicity, No. (%)** |  |  |  |  |  |  |
| NHW | 3374 (59.2) | 2913 (63.6) | 461 (41.2) | 180 (35.9) | 68 (29.4) | 14 (32.6) |
| NHAA | 981 (17.2) | 563 (12.3) | 418 (37.4) | 218 (43.5) | 113 (48.9) | 22 (51.2) |
| Other Ethnicity | 486 (8.5) | 378 (8.3) | 108 (9.7) | 45 (9.0) | 14 (6.1) | 1 (2.3) |
| Unknown Ethnicity | 857 (15.0) | 725 (15.8) | 132 (11.8) | 58 (11.6) | 36 (15.6) | 6 (14.0) |
| **SES, mean (SD); No.** |  |  |  |  |  |  |
| < HS Education | 0.08 (0.07); 4593 | 0.08 (0.06); 3807 | 0.09 (0.08); 786 | 0.10 (0.09); 265 | 0.12 (0.10); 104 | 0.14 (0.11); 18 |
| Unemployed | 0.07 (0.04); 4593 | 0.07 (0.03); 3807 | 0.08 (0.05); 786 | 0.08 (0.05); 265 | 0.09 (0.05); 104 | 0.09 (0.03); 18 |
| Annual Income < FPL | 0.13 (0.12); 4592 | 0.12 (0.114); 3806 | 0.14 (0.13); 786 | 0.16 (0.14); 265 | 0.17 (0.15); 104 | 0.22 (0.17); 18 |
| Persons per Mile<sup>2</sup> | 2650 (2340); 4593 | 2550 (2330); 3807 | 3150 (2310); 786 | 3600 (2580); 265 | 3780 (2500); 104 | 4490 (3220); 18 |
| **Comorbidity Score,**<br>**mean (SD); No.** | 2.53 (1.59); 4615 | 2.53 (1.58); 3838 | 2.53 (1.64); 777 | 3.02 (1.77); 272 | 2.78 (1.73); 107 | 3.89 (1.75); 18 |

Abbreviations: MM, Michigan Medicine; BMI, body mass index; NHW, non-Hispanic Whites; NHAA, non-Hispanic African Americans; SES, residential-level socioeconomic characteristics; HS, high school; FPL, federal poverty level.

**Figure 1.**
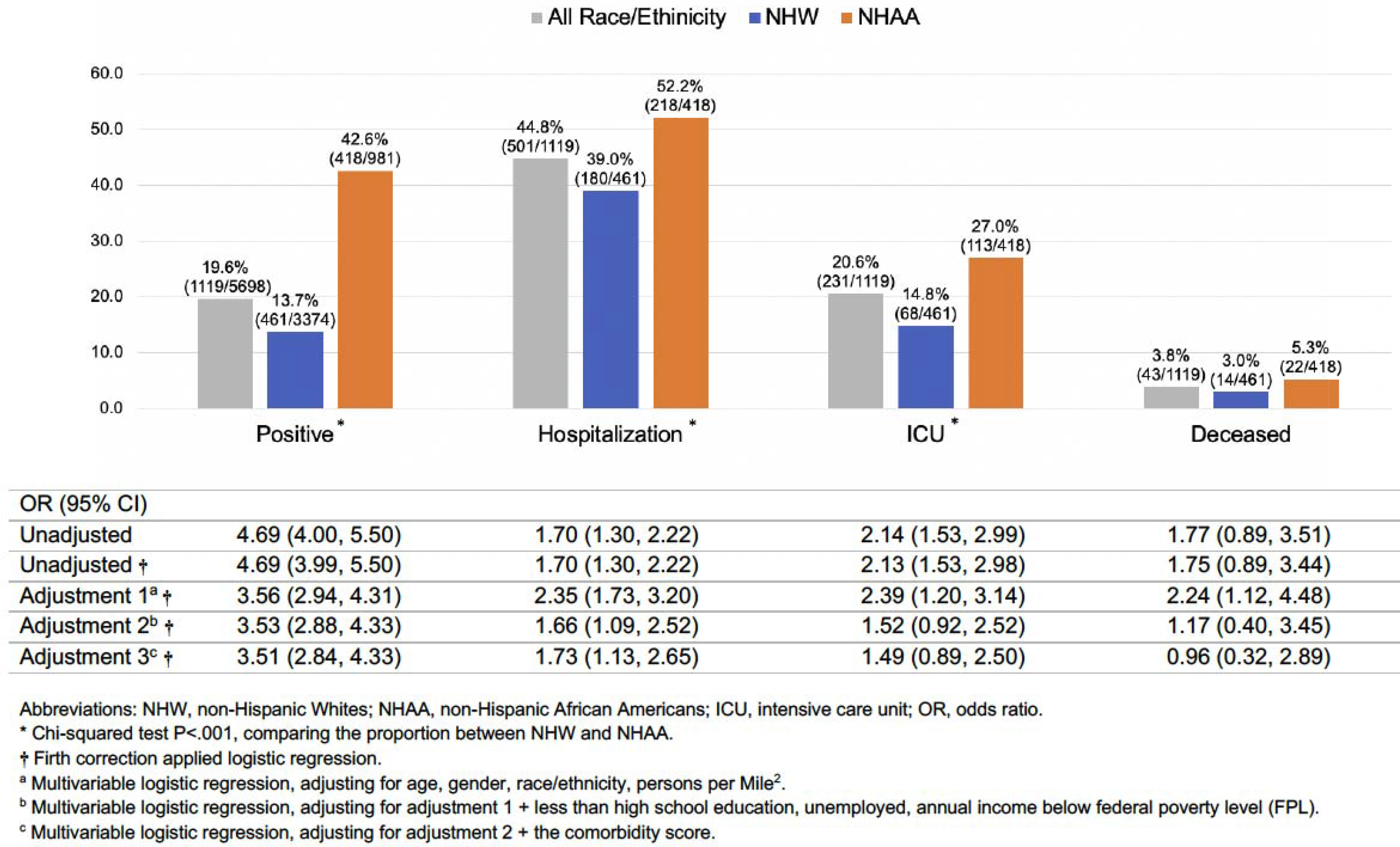
COVID-19 Outcomes by Race/Ethnicity.

### Association Analysis of COVID-19 Outcomes Using Multivariable Logistic Regression

#### Factors Associated with Getting Tested

Due to dependence on test availability, the testing guidelines varied during the time of the study. Comparing with the random controls from MM, overall, we noticed that male sex, current smoking, percentage of population with <HS education and percentage below FPL were inversely associated with the chance of getting tested whereas NHAA race/ethnicity, past smoking, age, BMI, and all comorbidities were positively associated with getting tested (eTable S5 in Supplement).

#### Factors Associated with being Positive or with COVID19 Susceptibility

##### Full cohort

We identified the following factors that differed between testing positive for COVID-19 and the randomly selected untested population from MM: NHAA vs NHW odds ratio (OR) was 7.11 (95% CI, 5.71-8.87; *P*<.001); older age (OR, 1.66 [95% CI, 1.52-1.82]; *P*<.001), higher BMI (OR,□ 1.35 [95% CI, 1.23-1.47]; *P*<.001), and alcohol consumption (ever vs. never; OR,□1.42 [95% CI, 1.17-1.73]; *P*<.001) (**Table 2**). Current smoking (self-reported in the latest EHR record) was observed as being protective against COVID-19 outcomes (OR, 0.34 [95% CI, 0.22-0.51]; *P*<.001), with a positive but insignificant association with past smoking (possibly due to quitting smoking for health reasons). In addition, population density was associated with testing positive (OR, 1.46 [95% CI, 1.33, 1.60]; *P*<.001). Percentage with income below FPL showed consistent association in protective direction, perhaps due to high correlation and overlap with other SES variables. **Table 2** indicates that pre-existing comorbidities were associated with testing positive. Conversely, a comparison between those **who testing positive versus those testing negative** showed *protective effects of existing comorbidity conditions*, a reflection of the bias due to targeted testing criteria.

**Table 2.** Multivariable Logistic Regression Comparing COVID-19 Positive Patients with Untested Controls and Tested Negatives.

|  | Positive (1) versus Control (0) |  |  | Positive (1) vs. Negative (0) |
| --- | --- | --- | --- | --- |
|  | Unmatched Control | Matched Control |  |  |
|  | All<br>(n<sub>0</sub>=5942, n<sub>1</sub>=766) | NHW<br>(n<sub>0</sub>=8521, n<sub>1</sub>=371) | NHAA<br>(n<sub>0</sub>=1029, n<sub>1</sub>=271) | All<br>(n<sub>0</sub>=3533, n<sub>1</sub>=766) |
|  | OR (95% CI) |  |  |  |
| Age* (y) | 1.66 (1.52, 1.82) | 1.43 (1.28, 1.6) | 1.75 (1.51, 2.02) | 1.37 (1.26, 1.49) |
| <50 | REF | REF | REF | REF |
| [50, 65) | 2.20 (1.80, 2.68) | 2.13 (1.65, 2.73) | 2.48 (1.79, 3.42) | 1.80 (1.48, 2.19) |
| [65, 80) | 1.88 (1.50, 2.37) | 1.55 (1.16, 1.07) | 2.19 (1.49, 3.23) | 1.65 (1.31, 2.08) |
| >=80 | 2.21 (1.59, 3.06) | 1.50 (1.02, 2.22) | 2.08 (1.10, 3.96) | 2.06 (1.48, 2.86) |
| Male Sex | 0.91 (0.77, 1.08) | 1.45 (1.18, 1.79) | 1.27 (0.96, 1.67) | 1.5 (1.27, 1.77) |
| BMI* | 1.35 (1.23, 1.47) | 1.34 (1.21, 1.49) | 1.33 (1.14, 1.54) | 1.24 (1.14, 1.35) |
| <18.5 | 0.67 (0.28, 1.57) | 0.77 (0.26, 2.30) | 1.47 (0.33, 6.49) | 0.705 (0.30,1.66) |
| [18.5, 25) | REF | REF | REF | REF |
| [25, 30) | 1.75 (1.36, 2.25) | 1.80 (1.34, 2.42) | 3.16 (1.71, 5.84) | 1.41 (1.10, 1.81) |
| >=30 | 2.04 (1.60, 2.60) | 2.20 (1.64, 2.94) | 6.15 (3.50, 10.82) | 1.63 (1.29, 2.06) |
| Ever-Smoker | 0.87 (0.71, 1.05) | 1.19 (0.95, 1.5) | 1.24 (0.91, 1.68) | 0.69 (0.57, 0.83) |
| Never-Smoker | REF | REF | REF | REF |
| Past-Smoker | 1.12 (0.91, 1.38) | 1.54 (1.22, 1.95) | 1.74 (1.25, 2.42) | 0.81 (0.66, 0.98) |
| Current-Smoker | 0.34 (0.22, 0.51) | 0.36 (0.2, 0.65) | 0.45 (0.24, 0.84) | 0.34 (0.22, 0.51) |
| Alcohol Consumption | 1.42 (1.16, 1.73) | 1.65 (1.28, 2.12) | 1.96 (1.41, 2.71) | 0.98 (0.81, 1.19) |
| Race/Ethnicity |  |  |  |  |
| NHW | REF | NA | NA | REF |
| NHAA | 7.11 (5.71, 8.87) | NA | NA | 3.53 (2.88, 4.33) |
| Other | 1.69 (1.28, 2.22) | NA | NA | 1.69 (1.29, 2.22) |
| Unknown | 0.16 (0.12, 0.22) | NA | NA | 0.86 (0.61, 1.21) |
| SES |  |  |  |  |
| < HS Education | 0.93 (0.83, 1.05) | 0.85 (0.71, 1.01) | 1.11 (0.94, 1.31) | 1.08 (0.96, 1.21) |
| Unemployed | 1.16 (1.04, 1.31) | 1.01 (0.86, 1.19) | 1.16 (1, 1.34) | 1.2 (1.07, 1.33) |
| Annual Income < FPL | 0.67 (0.59, 0.76) | 0.75 (0.64, 0.89) | 0.64 (0.53, 0.77) | 0.75 (0.65, 0.85) |
| Persons per Mile<sup>2</sup> | 1.46 (1.33, 1.60) | 1.48 (1.31, 1.66) | 1.17 (1, 1.37) | 1.28 (1.16, 1.41) |
| Comorbidity Score* | 1.87 (1.71, 2.05) | 2.09 (1.91, 2.29) | 2.43 (2.08, 2.82) | 0.81 (0.74, 0.88) |
| Respiratory | 3.28 (2.71, 3.97) | 4.00 (3.12, 5.14) | 3.46 (2.51, 4.77) | 0.69 (0.57, 0.85) |
| Circulatory | 2.46 (2.02, 2.99) | 3.15 (2.49, 3.98) | 4.86 (3.47, 6.79) | 0.62 (0.5, 0.76) |
| Any Cancer | 1.18 (0.95, 1.45) | 1.78 (1.41, 2.25) | 2.65 (1.88, 3.72) | 0.75 (0.61, 0.91) |
| Type II Diabetes | 1.92 (1.53, 2.41) | 3.07 (2.36, 4) | 3.05 (2.2, 4.22) | 0.97 (0.79, 1.2) |
| Kidney | 2.58 (1.98, 3.37) | 4.28 (3.19, 5.75) | 3.08 (2.13, 4.46) | 0.55 (0.44, 0.69) |
| Liver | 3.02 (2.18, 4.17) | 4.56 (3.18, 6.53) | 5.78 (3.07, 10.9) | 0.76 (0.58, 1.01) |
| Autoimmune | 2.55 (2.02, 3.21) | 3.84 (2.96, 4.96) | 3.42 (2.36, 4.95) | 0.95 (0.78, 1.17) |

Abbreviations: n<sub>1</sub>, sample size for COVID positive patients; n<sub>0</sub>, sample size for comparative group; OR, odds ratio; NHW, non-Hispanic Whites; NHAA, non-Hispanic African Americans; BMI, body mass index; NA, not applicable; REF, reference group; SES, residential-level socioeconomic characteristics; HS, high school; FPL, federal poverty level.
\*Odds ratios associated with one standard deviation (SD) change, with SD defined as the SD of the sample under consideration in each model.

##### Race/ethnicity-stratified

Former smokers were strongly associated with testing positive in both NHW (OR, 1.54 [95% CI, 1.22-1.95]; *P*<.001) and NHAA (OR, 1.74 [95% CI, 1.25-2.42]; *P*=.001). Overweight/obesity and having any cancer showed stronger effect in the race-stratified analysis, especially obesity in NHAA (OR, 6.15 [95% CI, 3.50-10.82]; *P*<.001). The stronger association was most likely due to stratification which addresses potential presence of interaction by race. All comorbidities and SES variables show similar directionalities as the full cohort.

#### Factors Associated with Prognosis among COVID-19 Diagnosed Patients

##### Full Cohort

Among the COVID-19 positive cohort, NHAA were 1.66 (95% CI, 1.09-2.52; *P*=.02) times more likely to be hospitalized, 1.52 (95% CI, 0.92-2.52; *P*=.1) times more likely of ICU admission, and 1.17 (95% CI, 0.4-3.45; *P*=.77) times more likely of death than NHW (**Table 3**), shown in Figure 1. Higher population density was associated with higher chance of hospitalization (OR, 1.27 [95% CI, 1.20-1.76]; *P*=.02), whereas older age, male sex and obesity consistently showed strong association with poor prognosis (**Table 3**). Type II diabetes (OR, 1.62 [95% CI 1.09-2.41]; *P*=.02) and kidney disease (OR, 2.55 [95% CI, 1.62-4.02]; *P*<.001) stood out as the strongest risk factors of hospitalization amongst the seven comorbidities. In spite of limited power due to sample size when comparing deceased versus alive, composite comorbidity score (OR, 1.72 [95%CI, 1.08-2.75]; *P*=.02) and kidney disease (OR, 2.82 [95%CI, 1.09-7.28]; *P*=.03) appear to be the strongest risk factors, in addition to age and male sex.

**Table 3.**
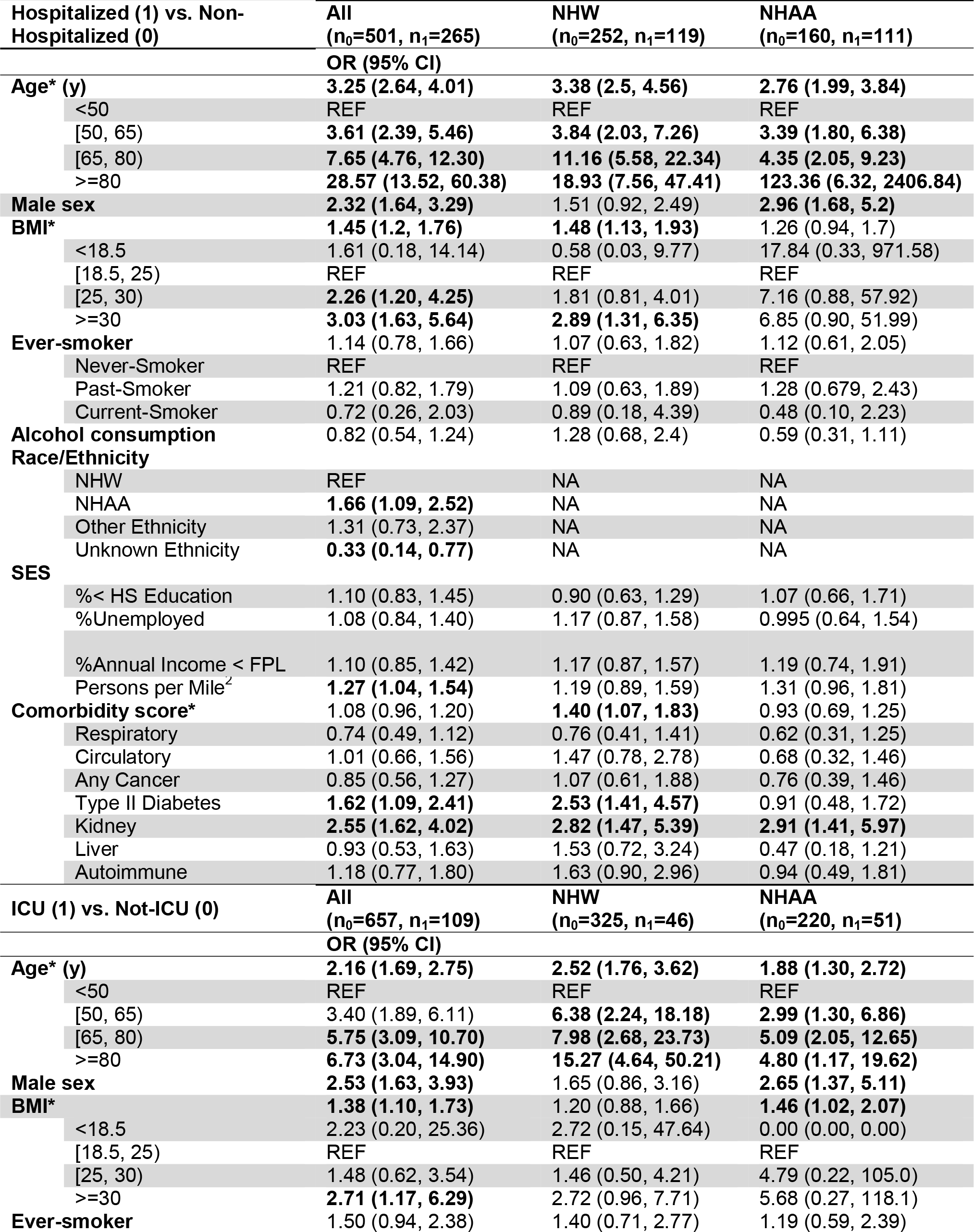
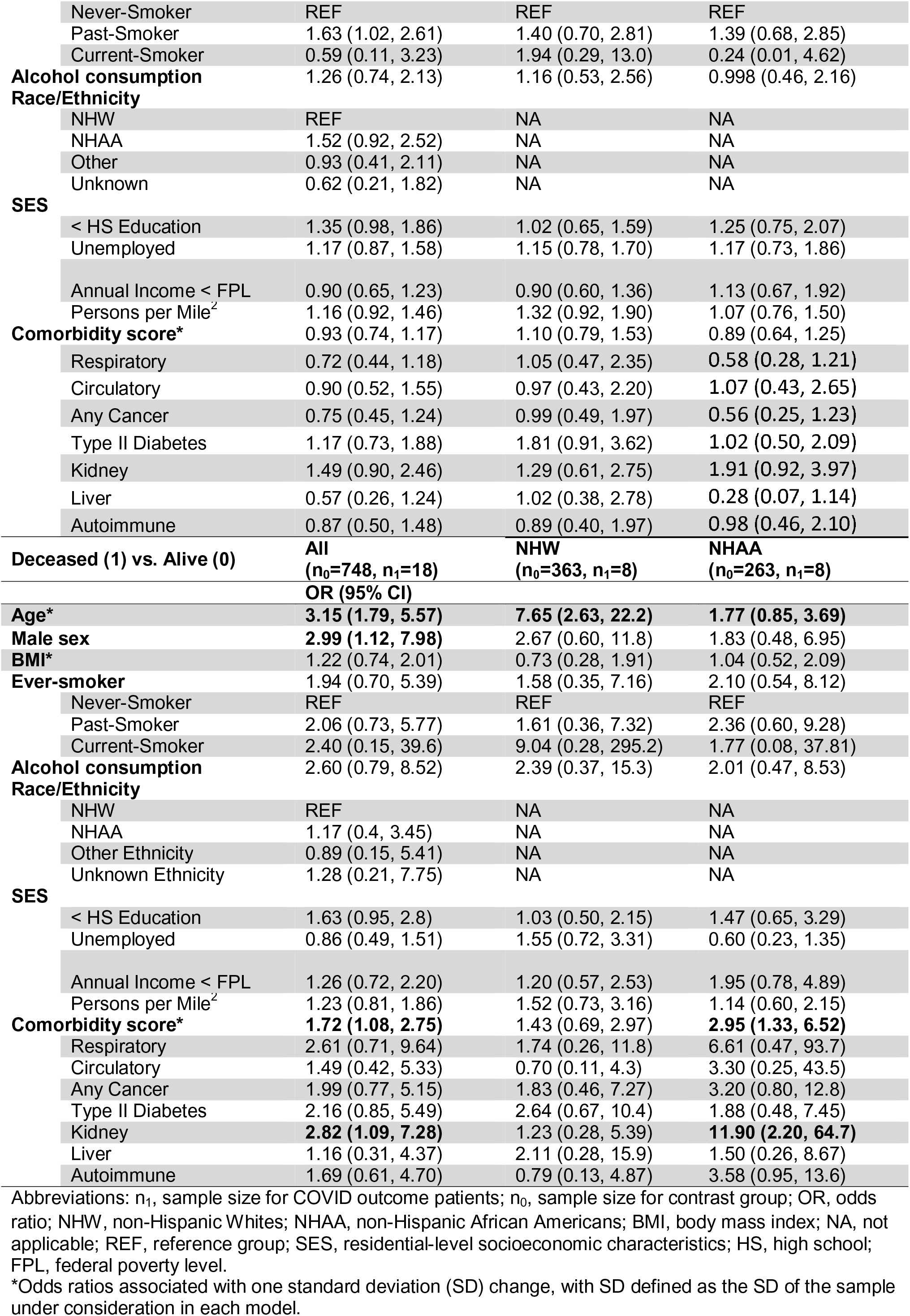
Multivariable Logistic Regression Odds Ratio of Prognostic Outcomes Among Diagnosed Patients.

##### Race/ethnicity-stratified

In NHW, patients with obesity and type II diabetes were 2.89 (95% CI, 1.31-6.35; *P*=.009) and 2.53 (95% CI, 1.41-4.57; *P*=.002) times more likely to be hospitalized compared to those without, respectively. In NHAA, males were at higher risk of being hospitalized (OR, 2.96 [95% CI, 1.68-5.2]; *P*<.001) than females, as well as being admitted to ICU (OR, 2.65 [95% CI, 1.37-5.11]; *P*=.004). In general, in addition to older age, patients with pre-existing kidney diseases had worse prognosis, especially for mortality in NHAA (OR, 11.9 [95% CI, 2.2-43.5]; *P*=.004).

## Discussion

This is one of the first studies to compare patient neighborhood SES, sociodemographic factors and health conditions from EHR data to uncover potential risk factors for observed racial disparities in COVID-19 susceptibility and prognosis. We identified racial and socioeconomic disparity in all COVID-19 outcomes (statistically significant for testing positive, hospitalization and suggestive for ICU admission and mortality due to small sample size). Pre-existing health conditions, and higher comorbidity burden were associated with poor prognosis.

This work contributes to a new area of COVID-19 research that incorporates both EHR data and SES data to rigorously examine racial/ethnic differences in disease susceptibility and prognosis. Other novel contributions include: (i) we compare the COVID-19 tested positive population with a random subset of the MM population to avoid the biased sample of who gets tested for COVID-19. (ii) We consider outcomes related to both susceptibility and prognosis. (iii) While most of the early studies present unadjusted analysis of various risk factors, we present a comprehensive analytic framework that attempts to adjust for an expanded set of potential confounders with suitably chosen comparison groups.

In general, our findings are consistent with existing studies. The observed high prevalence of kidney disease in COVID-19 mortality in NHAA can be possibly explained by the racial/ethnic differences in kidney disease^32^; males were at a higher risk of hospitalization and death, especially among those 50 years and older^33^; health conditions such as obesity, cancer, diabetes mellitus and renal conditions were prevalent in severe prognosis of COVID-19^22,34–36^. Notably, our findings largely agree with another recently published work looking at racial/ethnic differences in COVID-19 outcomes in an integrated-delivery health system in Louisiana^13^, which did not identify type II diabetes and kidney disease as risk factors of hospitalization as we do. Similar directional results but different strength of association with SES variables is likely because we used a continuous metric as opposed to the categorical measures used in Price-Haywood et al.^13^

Due to the prioritized testing scenario, there may be many asymptomatic or mild-symptomatic patients in the control population. Therefore, the comparison results between the positive and the unmatched control reveals the fact that in general, people with low SES and pre-existing health conditions had higher risk of developing severe disease outcomes after being infected with COVID-19. In contrast, a naïve comparison between the positive and the negative in the tested population leads to counterintuitive findings (protective effect of smoking, medication use, and worse comorbidity condition) contradicting findings in other COVID-19 studies^27,28^. This amplifies the need for choosing an appropriate control group.

This study has several limitations. First, we may not have captured all hospitalized patients given that only about half (48.3%) of the positive cohort had primary care at MM. It is possible that some of the “not hospitalized” patients actually were hospitalized elsewhere. Second, we did not consider the transfer patients from other hospitals as a special sub-group, who often had more severe outcomes. Third, early in COVID-19 course at MM, all COVID-19 patients were placed in regional infectious containment unit (RICU), some of which did not require ICU-level care. We suggest future study to define ICU patients using requirement of mechanical ventilators. Fourth, one may argue that the untested MM controls are intrinsically different than the tested cohort and do not serve as proper controls and may impact the estimation of the ORs observed in the susceptibility models. In the future, when testing is abundantly available, we may be able to restrict the patient population only to the catchment of MM or those who seek primary care at MM and exclude transfer patients. However, this is only a limitation in susceptibility models, and, the prognosis-outcome models only focused on the tested positive cohort and did not use the untested controls and thus, are not subject to the same selection issues. Lastly, the study results should be interpreted cautiously for generalizability given the demographics varies by region and by countries.

## Conclusion

The COVID-19 pandemic has reached every corner of the globe affecting millions and tens of millions of their loved ones. Our findings highlight that COVID-19 disproportionately affects those who are vulnerable: the elderly, those with pre-existing conditions, and those in population dense communities. Specifically, NHAAs have a greater risk burden – more likely to test positive, more likely to be hospitalized and more likely to be admitted to the ICU. Our results support targeted screening for the elderly and those with type II diabetes and kidney disease. Moreover, we call for increased investments in testing and prevention efforts in low SES, densely populated communities and racially diverse communities. It is these same communities that employ a greater proportion of essential workers who have been rightly called heroes but have still not been treated and protected as such. Genetics could also play a role in determining susceptibility and prognosis. Thus, it is important to expand this study to consider the multifactorial interplay of genetics and environment in the future.

## Data Availability

Data cannot be shared publicly due to patient confidentiality. The data underlying the results presented in the study are available from University of Michigan Medical School Central Biorepository for researchers who meet the criteria for access to confidential data.

https://research.medicine.umich.edu/our-units/central-biorepository/get-access

